# Twelve-Month Outcomes of Faricimab for Patients With Sub-optimally Responsive Diabetic Macular Oedema in a UK Tertiary Referral Centre

**DOI:** 10.1101/2024.12.13.24318978

**Authors:** Kamal El-Badawi, Benjamin Scrivens, Oluwaniyi Eke, Rehab Ismail, Lina Kobayter, Serena Salvatore

**Author notes:** Correspondence: Kamal El-Badawi, 10 Gainsborough Drive, Maidstone, Kent, ME16 0UZ.

## Abstract

**Purpose:** Evaluating 12-month visual and anatomical outcomes after switching to faricimab in diabetic macular oedema (DMO) patients with sub-optimal response to aflibercept 2mg.

**Patients and methods:** Sixty-two eyes of fifty patients were enrolled in this retrospective study at a UK tertiary referral centre. This consisted of DMO patients with sub-optimal response to aflibercept 2mg who were switched to faricimab. Four loading injections of faricimab were given at monthly intervals, followed by a treat-and-extend regime. The sub-optimal response was defined as CST >325 microns or >20% from best CST despite aflibercept 2mg at less than or equal to 8 weekly intervals(≤q8w) having completed a loading dose of aflibercept 2 mg (6 injections at monthly intervals). Outcome measures were best-recorded visual acuity (BRVA), central subfield thickness (CST), and treatment intervals.

**Results:** Baseline BRVA was 67.6 (±11.8) letters and CST was 406.4 (±105.9) µm, and the mean treatment interval was 6.5 (±1.8) weeks. At baseline, 24.2% of eyes were treated every 4 weeks (q4w), 19.4% every 6 weeks (q6w), and 56.5% every 8 weeks (q8w). After the 4^th^ faricimab loading dose, 54 patients continued on treat-and-extend faricimab. BRVA improved to 70.4 (±12.7) letters (p=0.007) while CST reduced to 372.8 (±132.0)µm (p=0.070). The mean injection interval improved to 7.4 (±2.6), 95%CI[0.1, 2.9]weeks. Subjects were followed up for 57.1 (±19.7) weeks, with a mean of 7.92 (±2.53) faricimab injections. At the latest follow-up, BRVA was stable at 68.7 (±14.6)(p=0.918) letters. CST had reduced further to 343.1 (±117.5) µm(p=0.034). Treatment intervals at the latest follow-up were: 3.2% q4w, 9.7% q6w, 30.6% q8w, 3.2% q10w, 11.3% q12w, 1.6% q14w, 6.5% q16w, with 53.2% on ≥q8w. The mean injection interval had also improved to 9.2 (±3.1) weeks(p=0.122).

**Conclusion:** In this study, DMO patients with sub-optimal response to aflibercept 2mg experienced improved anatomical outcomes and extended treatment intervals while maintaining vision by switching to faricimab.

## Introduction

Diabetic retinopathy (DR) is a frequent microvascular complication of diabetes mellitus (DM) that can result in macular thickening due to retinal vascular hyperpermeability, a condition known as diabetic macular oedema (DMO). DMO is the most common cause of moderate visual impairment in patients with DR.^1^ The global prevalence of DMO is estimated to be around 6.8%, equating to about 27 million adults worldwide being affected by DMO.^2^ The number of adults with DMO worldwide is anticipated to increase by 51.9% in the next 20 years.^3,4^

Intravitreal injection of anti-vascular endothelial growth factor (anti-VEGF) is currently the mainstay treatment of DMO.^5,6^ The landmark trials demonstrated improved anatomical and functional outcomes when followed up for at least two years.^7-10^

However, a subset of patients do not respond satisfactorily to anti-VEGF injections. A post hoc analysis of Diabetic Retinopathy Clinical Research (DRCR) Retina network Protocol T showed that 44% of patients receiving aflibercept 2mg for DMO had persistent macular oedema after two years of treatment.^11,12^ Additionally, recent studies examining real-world data have shown that real-world outcomes do not always reach the heights attained in clinical trials.^13^ Some patients have exhibited an inadequate response even with frequent and regular anti-VEGF therapy.^14,15^

This resistance phenomenon is believed to stem from a complex interplay of genetics, tachyphylaxis, and alternative angiogenic pathways beyond VEGF. Thus, various pathogenic mechanisms are implicated in DMO progression.^16^ Persistent DMO results in the propagation of oedematous changes driven by multiple inflammatory mediators earlier in the cascade, including angiopoietin-2 (Ang-2), which contributes to vascular destabilisation, vascular permeability, and neovascularization.^17-19^ The accumulation of reactive oxygen species (ROS) further exacerbates this process.

Faricimab (VabysmoTM, Roche/Genentech, Basel, Switzerland) is the first and currently the only bispecific molecule that targets VEGF and Angiopoietin-2 (Ang-2). Blocking the Ang-2 pathway improves vascular stability, culminating in DMO reduction.^20^ We aim to assess the real-world outcomes of intravitreal faricimab in DMO with sub-optimal response to previous treatment with aflibercept 2mg.

## Materials and Methods

### Study Design and Population

This retrospective study was conducted in a single tertiary centre: Bristol Eye Hospital, University Hospitals Bristol and Weston, Bristol, United Kingdom (UK). The study included 62 eyes of 50 patients with sub-optimally responsive DMO that were switched to faricimab. A sub-optimal response was defined after at least six monthly aflibercept 2mg doses, as:

- a central subfield thickness (CST) greater than 325 µm after the loading dose
- an increase of more than 20% from their best CST was recorded despite treatment intervals of q8w or less.

Exclusion criteria for the study included: 1) less than 6 doses of aflibercept at monthly intervals, 2) less than four doses of monthly faricimab injection, 3) intravitreal dexamethasone implant in the previous six months, 4) fluocinolone acetonide intravitreal implant in the previous three years, 5) macular laser within the previous 3 months to commencing faricimab. If an eye received an intravitreal steroid implant or macular laser, they were excluded from the study at that point.

Data collected from our Electronic Medical Records (EMR) included demographics, diabetes type, diabetic retinopathy (DR) grading, HbA1c, and treatment intervals.

Outcome measures were best recorded visual acuity (BRVA) in early treatment diabetic retinopathy study (ETDRS) letters, central subfield thickness (CST) measured using optical coherence tomography (OCT) (Topcon DRI OCT Triton plus; Tokyo, Japan) and treatment intervals in weeks. These were recorded at baseline (day of first faricimab injection), after loading and at the latest clinic visit.

### Treatment Protocol

Four doses of faricimab at 4-weekly intervals were given, with a review after the fourth loading dose. From this point, they moved on to the T&E regime, with intervals being maintained, extended, or reduced guided by anatomical outcomes compared to a reference CST value. The reference CST was the lowest CST recorded after loading (aflibercept 2mg or faricimab). Treatment intervals were reduced by 4 weeks if CST increased by >20% of the reference CST, maintained if CST increased by 10-20% of the reference CST, or extended by 4 weeks if CST increased by <10% of this reference value. This local treatment protocol was adapted and modified from the Personalised Treatment Interval (PTI) arm of the landmark trials in YOSEMITE and RHINE.^21^

### Statistical Analysis

Data was analysed using GraphPad Prism and Microsoft Excel. Mean averages were presented with standard deviation. A two-tailed paired t-test was performed, and a *p-*value less than 0.05 was considered clinically statistically significant.

## Results and discussion

### Demographics (Table 1)

The mean age of participants was 63.9 (±11.4) years, with the majority being male (56%) and predominantly of white ethnicity (80%). The mean HbA1c within 6 months of baseline was 64.5 (±18.2) mmol/mol.

**Table 1:** Demographic data from the Electronic Medical Records (EMR), 50 patients/62 eyes.

| Demographics |  |  |
| --- | --- | --- |
| Sex | Male | 28 (56%) |
|  | Female | 22 (44%) |
| Age | Mean | 63.9 (±11.4) |
| Ethnicity | White | 40 (80%) |
|  | Unknown/Not listed | 9 (18%) |
|  | White and black Caribbean | 1 (2%) |
| Type of diabetes | Type 1 | 7 (14%) |
|  | Type 2 | 43 (86%) |
| Grade of diabetic retinopathy (DR) | Mild non-proliferative (NP) DR | 15 (24%) |
|  | Moderate to severe NPDR | 25 (40%) |
|  | Proliferative DR | 22 (36%) |

### Baseline (n=62)

The mean number of injections received before the switch to faricimab was 17.3 (±10.7). All patients had received aflibercept 2mg before the switch to faricimab and six patients (9.7%) had initially been treated with ranibizumab followed by aflibercept 2mg before being switched to faricimab. Treatment received prior to faricimab initiation can be found in Table 2. At baseline, the BRVA was 67.6 (±11.8) letters, and CST was 406.4 (±105.9) µm (Table 3).

**Table 2:** A summary of treatment received for all eyes prior to switching to faricimab. All previous treatments were given in line with the study’s inclusion criteria

| Previous treatment history |  |  |  |
| --- | --- | --- | --- |
| Total number of previous injections |  | 1069 |  |
| Mean duration of previous DMO management (months) | | 62.9 ($\pm 36.4$) | |
| Mean number of previous injections | | 17.3 ($\pm 10.7$) | |
| | Mean of previous Eylea (Aflibercept 2mg) (n=62) | 16.5 ($\pm 9.5$) | |
| Mean injection interval (weeks) | | 7.5 ($\pm 2.8$) | |
| Treatment intervals (n=62 eyes) | 4 weeks | 15 (24.2%) |  |
| | >4≤6 weeks | 12 (19.4%) | |
| | >6≤8 weeks | 35 (56.5%) | |
| Previous DMO management (n=62 eyes) | Anti-VEGF intravitreal injections | Eylea® (Aflibercept 2mg) | 62 (100%) |
|  |  | Lucentis® (Ranibizumab 0.5mg) | 6 (9.7%) |
|  | Intravitreal dexamethasone implant (Ozurdex®) |  | 4 (6.5%) |
|  | Intravitreal fluocinolone acetonide implant (Iluvien®) |  | 2 (3.2%) |
|  | Macular Laser |  | 16 (25.8%) |
|  | Vitrectomy |  | 6 (9.7%) |

**Table 3:**
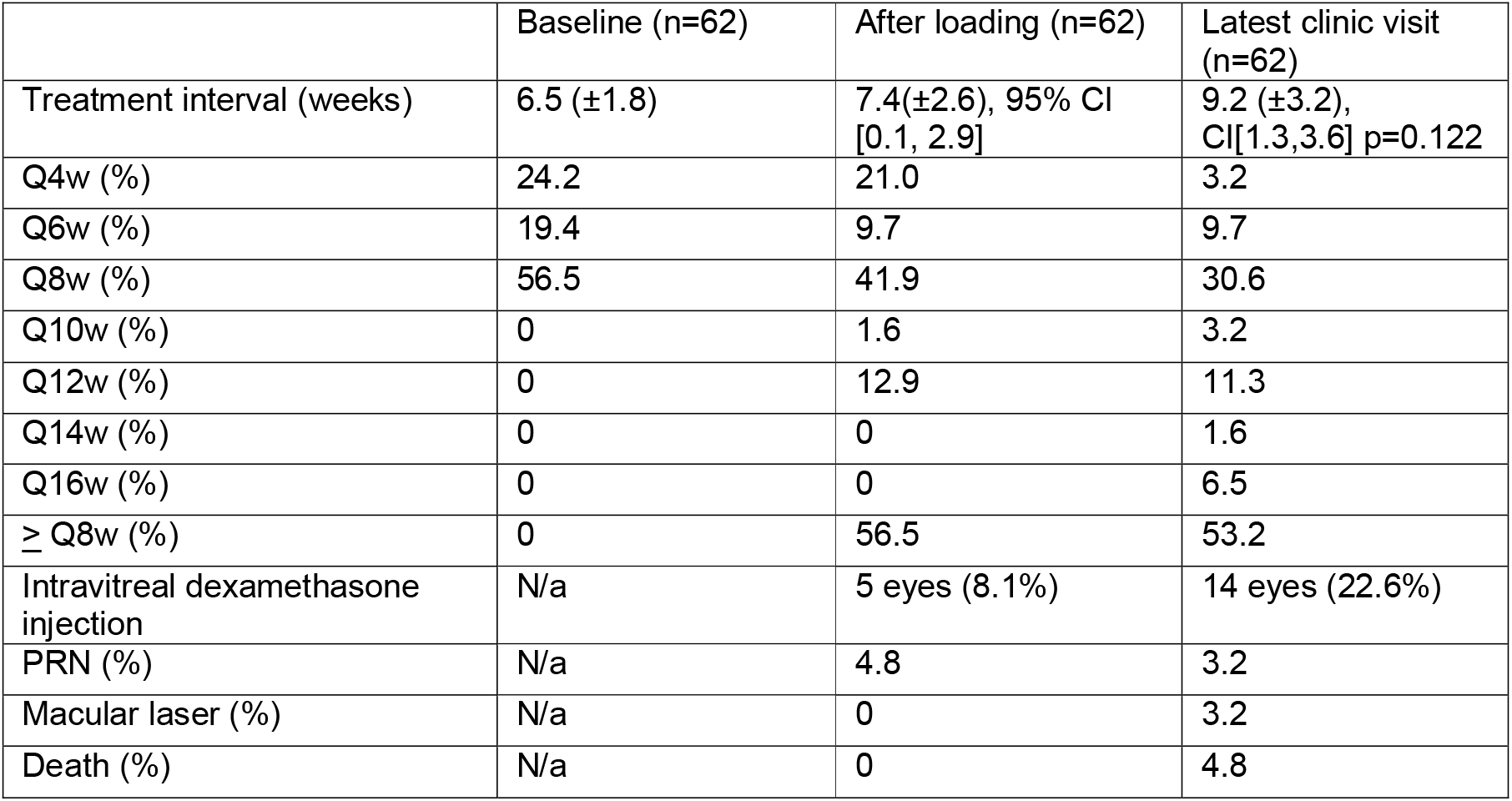
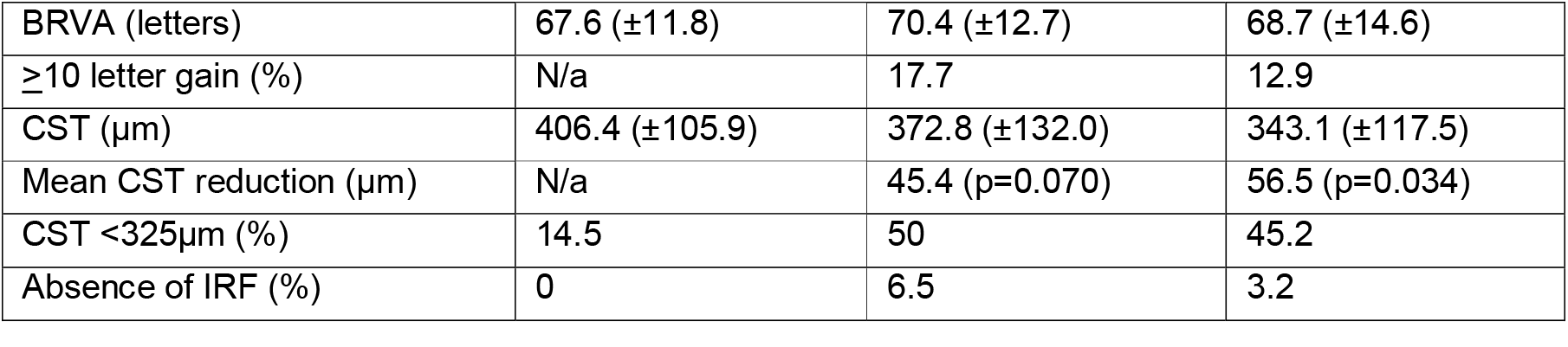
Summary of visual outcomes, anatomical outcomes and treatment intervals at baseline, post-loading, and latest clinic visit.

### Visit after Loading

After the fourth injection in the loading phase, the mean review time was 6.5 (±1.9) weeks. Fifty-four eyes continued in the T&E pathway as per the local protocol; 5 eyes were switched to an intravitreal dexamethasone implant due to poor response, and three were switched to a pro re nata (PRN) treatment regime and therefore excluded from further analysis (Table 3).

BRVA was 70.4 (±12.7) letters, with a gain of 2.8 letters, 95% CI [0.8, 4.7] letters from baseline (p=0.007). 11 eyes (17.7%) gained ≥10 letters. CST improved to 372.8 (±132.0) µm, 95% CI [9.7, 81.0] (p=0.070). At this point, 31 eyes (50%) had CST <325 µm, 4 eyes (6.5 %) had no IRF at this visit compared to 0% at baseline. The treatment intervals prescribed from this visit were as follows: 13 eyes (21%) q4w, 6 eyes (9.7%) q6w, 26 (41.9%) q8w, 1 eye (1.6%) q10w, and 8 eyes (12.9%) q12w. This equates to 56.5% being on treatment intervals of 8 weeks or more. The mean treatment interval was now 7.4 (±2.6) weeks, a gain from baseline of 0.9 weeks 95% CI [0.1, 2.9].

### Latest clinic visit outcomes

Forty-one eyes remained on the T&E pathway with faricimab; 14 switched to intravitreal dexamethasone implants; 2 patients (3 eyes) died, and one discontinued treatment (Table 3). The mean follow-up was 57.1 (±19.7) weeks, receiving a mean of 7.92 (±2.53) faricimab injections from baseline.

Outcome metrics were available for 42 eyes; at their latest clinic visit, the BRVA was stable at 68.7 (±14.6) letters (p=0.918). Eight (12.9%) gained ≥10 letters. CST was 343.1 (±117.5) µm with a reduction from baseline of 56.5µm, 95% CI [4.6,108.3] (p=0.034). Two eyes (3.2%) had resolution of diabetic macular oedema. Thirty-nine eyes (62.9%) had a reduction in CST compared to baseline. The treatment intervals recommended at the latest clinic visit were based on CST reference values. These are as follows: 2 eyes (3.2%) q4w, 6 eyes (9.7%) q6w, 19 eyes (30.6%) q8w, 2 eyes (3.2%) q10w, 7 eyes (11.3%) q12w, 1 eye (1.6%) 14 weeks q14w, 4 eyes (6.5%) 16 weeks (q16w). The mean injection interval was 9.2 (±3.2) weeks, a gain of 2.4 weeks, 95% CI[1.3,3.6] (p=0.122). 53.2% of those on the T&E regime at the latest clinic visit were on a treatment interval of 8 weeks or more.

A total of 14 eyes (22.6%) had poor response to treatment, demonstrated by either no improvement or a significant increase in CST, and these were switched to intravitreal dexamethasone implant. Two eyes (3.2%) received adjunctive treatment with macular laser and were subsequently excluded from the analysis at this point.

The mean HbA1c at the latest clinic appointment remained stable at 65.5 (±18.7), CI[3.9, −1.8], p=0.473mmol/mol.

### Safety

A total of 491 faricimab injections were analysed. In our study, 2 patients (3 eyes) died from pre-existing health conditions unrelated to their intravitreal treatment. One patient (2 eyes) had a flare-up of her pre-existing anterior uveitis and responded well to topical steroid treatment only. There were no other safety concerns reported.

## Discussion

This study evaluated faricimab’s effectiveness in treating DMO in patients who showed sub-optimal response to prior aflibercept 2mg therapy. Switching this cohort to faricimab allowed us to extend treatment intervals, enhance macular anatomy, and maintain stable vision outcomes.

The cohort’s demographic profile and significant retinal disease aligns with DMO epidemiology. These challenging cases had a high treatment burden, averaging 17.3 aflibercept 2mg injections prior to the switch. A small subset remained poorly responsive and required intraocular steroid implants post-switch.

Real-world studies corroborate faricimab’s benefits in similar challenging cohorts, summarised in Table 4.^22-28^ Like ours, these studies demonstrate stable vision, anatomical improvements, and no new safety concerns following the switch. Visual acuity in our cohort was maintained throughout, with modest, non-significant improvements after the loading phase. Anatomical outcomes showed a significant CST reduction, with 67.7% achieving CST reduction after loading, increasing to 84.8% at the latest visit.

**Table 4:** Summary table of current literature on the use of faricimab in DMO.

|  | Eyes | Patients | Follow-up (months) | Number of injections | VA change | CST change | Treatment interval post-loading | Were eyes reloaded after switching to faricimab? | Clinical rationale for reloading after switching to faricimab | Additional comments |
| --- | --- | --- | --- | --- | --- | --- | --- | --- | --- | --- |
| **Rush 2022 (22)** | 24 | 24 | 4 | 3 | 5.0 | -59.6 | Not reported | Yes | As specified in the treatment protocol |  |
| **Rush 2023 (23)** | 52 | 51 | 12 | >6 | 6.5 | -59.6 | 39.2% reached >8 weeks after loading | Yes | As specified in the treatment protocol |  |
| **Bailey 2023 (24)** | 225 | Not reported | NA | * | 1.1 | Not reported | Last mean treatment interval 7.2 weeks | Unknown | Real-world retrospective medisoft data, across multiple sites with varying local protocols. Not specified which eyes were reloaded | *Between 5th and 6th injection shown |
| **Pichi 2024 (25)** | 100 | 100 | 6 | 2.9 | 5 | -67.9 | N/A | No | The protocol specified to switch to OCT-guided as-needed regime |  |
| **Quah 2024 (26)** | 72 | 58 | 2.5 | 4.7 | 2.5 | -104 | N/A | Yes | As specified in the treatment protocol |  |
| **Tatsumi 2024 (27)** | 29 | 21 | N/A | 4 | -3.3 | -53 | N/A | No | The protocol specified to switch to an OCT-guided and clinician-discretion as-needed regime |  |
| **Durrani 2024 (28)** | 69 | 53 | N/A | 3 | -1 | -57 | 25 eyes were able to be extended by 2 or more weeks | No | Clinician discretion, guided by OCT |  |
| **Borchet 2024 (29)** | 44 | 28 | 6 | 5 | 1 | -38 | Last treatment interval 6.6 weeks (1.4-week difference from baseline) | Yes** | The decision to reload was at the discretion of the clinician | **78% of DMO eyes were reloaded, the decision was made at clinician's discretion |
| **Present study, El-Badawi 2024** | 62 | 50 | 13 | 7.9 | 1.1 | -56.5 | The last treatment interval 9.2 weeks, a gain of 2.4 weeks from baseline | Yes | As specified in the treatment protocol |  |

**Abbreviations:** Ang-2, angiopoietin-2; Anti-VEGF, anti-vascular endothelial growth factor; BRVA: rest recorded visual acuity; CST, central subfield thickness; DM, diabetes mellitus; DMO, diabetic macular oedema; DR, diabetic retinopathy; ETDRS, early treatment diabetic retinopathy study; OCT, optical coherence tomography.

Compared to YOSEMITE and RHINE trials, where 78-86% achieved <325 µm CST by 52 weeks in the PTI group, 50% of our cohort achieved this after the loading phase, and 45.2% maintained it at the most recent visit. Similarly, our findings align with real-world data from Rush et al., where 37.5% achieved a CST of <300 µm after four months. Given the complexity of our treatment-experienced cohort, these outcomes are highly encouraging.

A key finding in our study was the extension of the injection interval to an average of 9.2 weeks at 12 months. This is particularly noteworthy given the significant psychological and economic burden that frequent injections impose on patients, their caregivers, and healthcare systems. These burdens, well-documented in the literature, can adversely impact adherence and overall quality of life^29,30^. In our study, the average injection interval increased by 2.4 weeks, with the proportion of patients needing injections every 8 weeks dropping from 56.5% to 30.6%. Notably, 53.2% of our cohort achieved intervals of 8 weeks or longer by 12 months, surpassing the 39.2% reported by Rush et al.^23^ Comparable results were seen for 12-week intervals, and fewer patients required 4-week injections.

While Durrani et al. demonstrated interval extension, their study lacked long-term follow-up and a defined loading protocol.^28^ Reducing the injection burden significantly benefits patients and caregivers by easing logistical and financial pressures. For example, this approach could free up 2,040 appointments and save £230,520 annually per 1,000 patients in appointment costs alone, not accounting for additional savings from transportation or lost work hours.

The psychological relief of extended intervals and fewer treatments is invaluable. As these data reflect only the first year with mandatory loading doses, further improvements in the second year could enhance long-term outcomes and healthcare efficiency.

No new safety signals were identified across the 491 injections administered. Two unrelated deaths and a case of anterior uveitis flare in a patient with a known history of the condition were reported. Importantly, no endophthalmitis, retinal vasculitis, or new intraocular inflammation were reported.

The variability in BRVA and CST outcomes highlights the need for predictive biomarkers to determine better which patients are likely to respond favourably to faricimab. Also, our cohort’s predominantly white demographic limits the generalisability of these findings to other ethnic groups. Additionally, the retrospective nature of this study imposes inherent limitations. Prospective, long-term, real-world studies will provide deeper insights into managing this challenging patient population.

## Conclusion

This study demonstrated that in DMO cases with sub-optimal response to aflibercept 2mg, switching to faricimab can improve anatomical outcomes, maintain visual acuity, and extend injection intervals. These benefits not only enhance patient outcomes but also reduce the treatment burden. This approach allows for better alignment with patient needs while improving efficiency and lowering healthcare costs.

## Data Availability

All data produced in the present study are available upon reasonable request to the authors

## Acknowledgments

We extend our heartfelt gratitude to the patients and their families for their participation and trust and to the clinical and administrative staff at the Bristol Eye Hospital for their invaluable support in patient care and data collection.

Special thanks go to Miss Salvatore and Dr. Kobayter for their efforts in developing and implementing the treatment protocol, which was instrumental in achieving this study’s outcomes. We are also grateful to Mr Eke for his valuable assistance with the writing and preparation of this manuscript.

## Disclosure

Kamal El-Badawi: No conflicts of interest to declare.

Benjamin Scrivens: Educational grant from Roche.

Oluwaniyi Eke: Travel bursary and advisory board Bayer, speaker fees Roche.

Rehab Ismail: No conflicts of interest to declare.

Serena Salvatore: Advisory board for Bayer, Roche; Speaker fees and travel bursary from Alimera

Sciences, Roche Products Limited, Bayer.

Lina Kobayter: Advisory board and travel bursary from Roche Products Limited.. Speaker fees from

Bayer, Alimera Sciences and Roche Products Limited.

